# Is mammography screening beneficial: An individual-based stochastic model for breast cancer incidence and mortality

**DOI:** 10.1101/2020.01.30.20019596

**Authors:** Thuy T. T. Le, Frederick R. Adler

**Affiliations:** DEPARTMENT OF MATHEMATICS, UNIVERSITY OF UTAH, 155 SOUTH 1400EAST, SALT LAKE CITY, UT, 84112, USA; DEPARTMENT OF HEALTH MANAGEMENT AND POLICY, SCHOOL OF PUBLIC HEALTH, UNIVERSITY OF MICHIGAN, 1415 WASHINGTON HEIGHTS, ANN ARBOR, MI 48109, USA

**Keywords:** individual-based model, stochastic model, mathematical model, breast cancer, mammography screening, overdiagnosis

## Abstract

**BACKGROUND:** The benefits of mammography screening have been controversial, with conflicting findings from various studies.

**METHODS:** We hypothesize that unmeasured heterogeneity in tumor aggressiveness underlies these conflicting results. Based on published data from the Canadian National Breast Screening Study (CNBSS), we develop and parameterize an individual-based mechanistic model for breast cancer incidence and mortality that tracks five stages of breast cancer progression and incorporates the effects of age on breast cancer incidence and all-cause mortality.

**RESULTS:** The model accurately reproduces the reported outcomes of the CNBSS. By varying parameters, we predict that the benefits of mammography depend on the effectiveness of cancer treatment and tumor.

**CONCLUSIONS:** In particular, patients with the most rapidly growing or potentially largest tumors have the highest benefit and least harm from the screening, with only a relatively small effect of age. However, the model predicts that confining mammography populations with a high risk of acquiring breast cancer increases the screening benefit only slightly compared with the full population.

## 1. Background

Breast cancer is one of three most commonly diagnosed cancers in women, making up 30% of all cancer cases in women in the United States in 2018 [1]. Screening mammography was introduced to detect small and more treatable tumors before they cause symptoms. Several trials, such as the Health Insurance Trial [2], the Edinburgh randomised trial [3,4], the Canadian National Breast Screening Study (CNBSS) [5] and the Swedish Two-Country Trial [6, 7], have quantified the benefits of screening mammography. The Swedish study and many others reported that breast cancer mortality was significantly reduced due to screening mammography [6, 8], while the CNBSS found no benefits [5, 9]. In addition, Welch et. al. also found no benefit in their analysis of the SEER data [10]. These contradictory conclusions have spurred intense debate over the benefits of screening mammography. The wide implementation of screening mammography has led to an increased rate of small tumor detection and a decreased rate of large tumor detection over the last decades [10]. The primary cost of screening is overdiagnosis of small benign or unaggressive tumors that would have remained asymptomatic during a patient’s lifetime, turning a healthy individual into a patient, and requiring follow-up tests and treatments with deleterious side effects including death [11]. Overdiagnosis also results in unwanted economic and psychological burdens. To address the controversy, the WISDOM study based on a woman’s individual risk was initiated in the United States in 2016 [12].

Studies based on statistical or stochastic models [13–15] have quantified the influence of various factors such as age, screening frequency and adherence behavior on the benefits and harmful effects of mammography screening based on different data sources or trials other than the CNBSS. Most of transition probabilities in these models were held constant or age-dependent, and thus did not include the effects of tumor heterogeneity across patients. Using the CNBSS, several analyses ([16, 17] and references therein) have estimated screening sensitivities, transition probabilities and sojourn time distributions. As far as we know, no study has developed a mathematical model to quantify the benefits and harm of screening that explicitly takes tumor heterogeneity into consideration. In this work, we propose a mechanistic model focusing on differences among individuals that provides a mathematical tool to gain insight into breast cancer progression. This model includes all possible transitions of breast cancer before and after detection from cancer incidence and detection through progression, treatment and mortality. Our central focus is on unmeasured heterogeneity, which we include through variation in the aggressiveness of tumor growth and maximum tumor size. By including unaggressive cancers, we are able to model the role of unmeasured heterogeneity in incidence levels, detected tumor sizes, and long-term outcomes to address the balance between costs and benefits of screening. The benefits are measured as the increase in 25-year survival. The costs are the increase in overdiagnosis quantified in two ways, through the difference in the number of patients diagnosed [18], and through the number who would have died due to other causes if treatment were relatively ineffective.

The proposed model is designed to first reproduce the cancer incidence and mortality in the CNBSS with a minimum of parameter fitting to the data itself [5]. By varying key model parameters, we simulate different scenarios of tumor aggressiveness and cancer treatment effectiveness to quantify their effect on the benefit and harm of mammography screening in a population.

The paper first presents the model framework and describes how parameters were estimated from the literature and the CNBSS. Because of the focus on unmeasured differences in underlying cancers, we term this the Breast Cancer Heterogeneity Aggressiveness Model (BCHAM). Using BCHAM, we experiment with the effectiveness of treatment and the underlying mean and variance of tumor aggressiveness to identify when mammography should provide the greatest benefit and the least harm.

## 2. Materials and Methods

### 2.1. The CNBSS

The CNBSS has been described in detail [5, 19], and we here summarize its key features (Figure 1a). The CNBSS was designed to investigate the benefits of mammography screening in women aged 40 − 59. The patients were followed up for up to 25 years (22 years on average). A population of 89, 835 healthy women aged 40 − 59 was randomly assigned to mammography (five annual mammography screens) and control (no mammography). Women in the mammography arm received both annual mammography and physical examination for the first 5 years of follow-up. In the control arm, women aged 40 − 49 received only a single physical examination at enrollment, and women aged 50 − 59 received annual physical examination for the first 5 years of follow-up. Participants were considered eligible if they were in good health, had no mammography in the previous 12 months, and had no history of breast cancer. The number of detected breast cancers, breast cancer mortality and all-cause mortality was recorded during the follow-up period.

**Figure 1.**
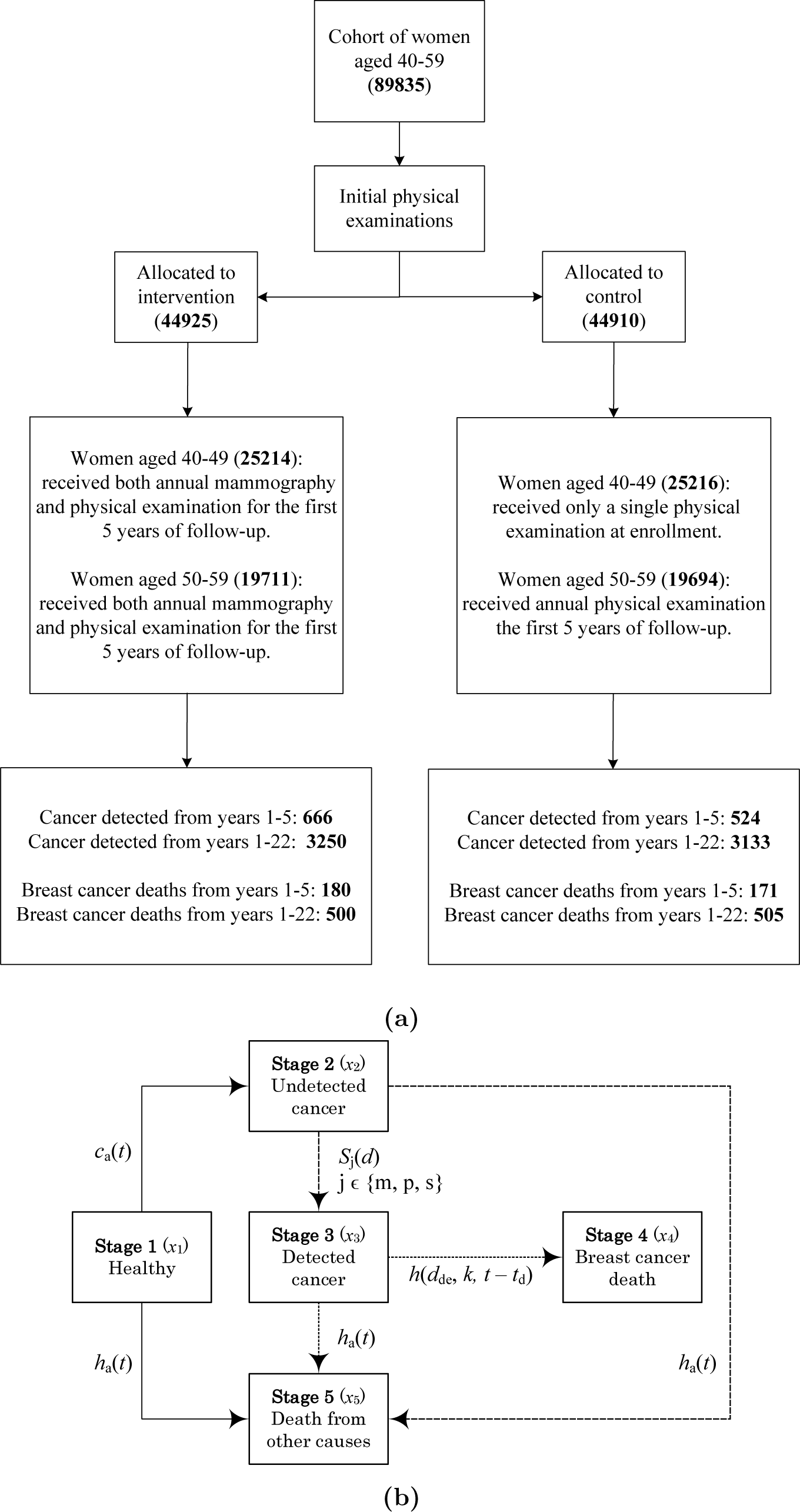
a) Flow diagram of the CNBSS [19]. Values in parentheses indicate the number of individuals in each compartment. b) Diagram of the stages of breast cancer incidence and mortality in BCHAM. Solid arrows indicate transitions from the healthy state, long-dashed arrows from the undetected state, and short-dashed arrows from the detected state

### 2.2. BCHAM: An individual-based stochastic model

We use a five-compartment model to track the number of women at each cancer stage via the probabilities and rates of transition between consecutive stages (Figure 1b). Let *a* denote the age of a woman at enrollment and *t* the time since the beginning of follow-up. The rate of cancer incidence, *c*_*a*_(*t*), is a bell-shaped function based on the report of Canadian Cancer Registry and Health Statistics Division [20]. The rate of non-breast cancer mortality is captured by an exponential function *h*_*a*_(*t*) obtained from the 1991 Canadian statistics reported in [21].

Several models of tumor growth have been used in the literature [22, 23]. We model tumor diameter at time *t* with initial diameter *d*_ini_ at initial time *t*_ini_, *d*(*t, t*_ini_, *d*_ini_, *k*), with a Gompertz model of human breast cancer growth [22]. The key parameter *k* is the tumor aggressiveness constant. Detection sensitivities of a tumor are modeled by sigmoid functions [24]. *S*_*m*_(*d*), *S*_*p*_(*d*) and *S*_*s*_(*d*) denote the tumor detection sensitivities of mammography, physical screening and self-examination respectively during the follow-up period. To capture undetected cancers entering the study, we let *S*_*b*_(*d*) be the tumor detection sensitivity of self-examination before the beginning of the study to eliminate candidates with noticeable tumors. Let *t*_d_ represent the time when a tumor is detected (the time when a patient moves from Stage 2 to 3). Suppose that breast cancers originate from a single cell of the diameter *d*_0_ at time *t*_0_, the time when a woman moves from Stage 1 to 2. Let *d*_de_ =*d*(*t*_d_, *t*_0_, *d*_0_, *k*) be the size of a tumor at detection time *t*_d_ > *t*_0_. The hazard of cancer mortality depends on the size at detection, tumor aggressiveness and time since detection according to *h*(*d*_*de*_, *k, t* − *t*_*d*_), which follows a two-parameter Weibull distribution based on the probability of cancer mortality [25]. This function includes a parameter *α* that captures the effectiveness of treatment. All parameter values are presented in Tables 1a and 1b.

**Table 1.** Model parameters and values.

| Parameters | Values | Units |
| --- | --- | --- |
| Cancer mortality hazard parameter $Q$ [26] | 0.0067 | 1/mm<sup>Z</sup> |
| Cancer mortality hazard parameter $Z$ (estimated) | 1.06 | |
| Cancer mortality hazard parameter $\omega$ [27] | 1.272 | |
| Scale factor in cancer incidence $\gamma$ (estimated) | 1.54 | |
| Maximum tumor diameter ( [22, 23]), $d_{max}$ | $\text{unif}(1, 128)^{(1)}$ | mm |
| Tumor diameter of a single cell, $d_0$ | 0.0124 | mm |
| Current tumor diameter at time $t$ with an initial diameter $d_0$ at time $t_0$, $d(t, t_0, d_0, k)$ [22] | $d_{max} \left( \frac{d_{min}}{d_{max}} \right)^{\exp(-12k(t-t_0))}$ | mm |
| Tumor aggressiveness, $k$ [22] | $\log(k) \in \mathcal{N}(\mu_k, \sigma_k)$ | month<sup>-1</sup> |
| Cancer incidence parameters $\mu_{age}, \sigma_{age}$ [20] | 76.86, 19.5 | years old |
| True cancer incidence rate, $c_a(t)$ [20] | $\frac{0.1967\gamma}{\sigma_{age}\sqrt{2\pi}} \exp\left(\frac{-(t+a-\mu_{age})^2}{2\sigma_{age}^2}\right)^{(2)}$ | year<sup>-1</sup> |
| Rate of other cause mortality, $h_a(t)$ [21] | $0.208 \times 10^{-5} \exp(0.1196(t+a))$ | year<sup>-1</sup> |
| Detection sensitivity of a tumor size $d$, $S_j(d)$, $j \in \{b, m, p, s\}$, [28] | $\frac{\exp((d-b_{j2})/b_1)}{1 + \exp((d-b_{j2})/b_1)}$ | |
| Hazard rate of cancer mortality $h(d_{de}, k, t)$, based on [26, 27] | $k^\alpha d_{de}^Z \omega t^{\omega-1}$,<br>$\alpha = \frac{\log(Q/15^\omega)}{\log(k_m)} = 3.19$ | |
| Detection sensitivity constants $b_1, b_{m2}, b_{p2}, b_{s2}$ [19, 28] | 1.5, 6.33, 18.5, 20 | mm |
| Detection sensitivity constants $b_{b2}$ (estimated) | 40 | mm |
| Mean of $\log(k)$ adapted from [22], $\mu_k$ | -2.9 | month<sup>-1</sup> |
| Standard deviation of $\log(k)$ [22], $\sigma_k$ | 0.71 | month<sup>-1</sup> |
| Mean of $k$, $k_m$ | $\exp(\mu_k + \sigma_k^2/2) = 0.0708$ | |

| Parameters | Values |
| --- | --- |
| Age of participants, $a$ | $\text{unif}(40, 59)$ |
| Average follow-up time, $t$ | 0 – 22 |
| Population size, $N_0$ | see Figure 1a |
| Mammography screening schedule for women aged 40 – 59 in the mammography arm, $T_m$ | $\{0^{\text{th}}, 1^{\text{st}}, 2^{\text{nd}}, 3^{\text{rd}}, 4^{\text{th}}\}$ |
| Physical examination schedule for women aged 40 – 59 in the mammography arm, $T_p$ | $\{0^{\text{th}}, 1^{\text{st}}, 2^{\text{nd}}, 3^{\text{rd}}, 4^{\text{th}}\}$ |
| Physical examination schedule for women aged 40 – 49, $T_p$ in the control arm | $\{0^{\text{th}}\}$ |
| Physical examination schedule for women aged 50 – 59, $T_p$ in the control arm | $\{0^{\text{th}}, 1^{\text{st}}, 2^{\text{nd}}, 3^{\text{rd}}, 4^{\text{th}}\}$ |

(b) Study-specific values of BCHAM model parameters based on the design of the CNBSS [5, 23].

At enrollment, the participants can be in either the healthy or the undetected cancer compartment. As time passes, they can transfer between stages (Fig. 1b).

Stage 1: An individual in the healthy compartment may develop undetected breast cancer at a rate *c*_*a*_(*t*) or die due to other causes apart from breast cancer at a rate *h*_*a*_(*t*) (solid arrows in Fig. 1b).

Stage 2: During follow-up, an individual with undetected cancer can be detected with a probability of *S*_*j*_(*d*), *j* ∈ {*m, p, s*}, or die of other causes at a rate of *h*_*a*_(*t*) (long-dashed arrows in Fig. 1b).

Stage 3: An individual with detected cancer may die of breast cancer at a rate of *h*(*d*_*de*_, *k, t* − *t*_*d*_) or of other causes at rate *h*_*a*_(*t*) (short-dashed arrows in Fig. 1b).

We simulate a population of *N*_0_ individuals. The total women in the *i*-th compartment at time *t* is 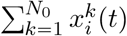 where 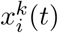, *i* = 1, …, 5 is an indicator function of the stage *i* of a individual *k* at time *t*.

### 2.3. Parameter calibration based on the CNBSS

We simulate the model using the CNBSS [5] to calibrate model parameters that cannot be estimated independently from the literature. In the CNBSS, the large number of breast cancers detected during the first year of follow-up (253 and 170 diagnosed cancers in the mammography and control arms respectively (Table 1 and [5]) suggests that at enrollment the participants may have been either in the healthy or undetected cancer compartment. To capture this, we began each simulated patient as healthy 6 years prior to the study initiation, with 6 years as a sufficient time period for previously originating cancers to be diagnosed before the first year of the study. In accordance with the study design, we assume that cancers can be diagnosed only by self physical examination with the detection sensitivity *S*_*b*_(*d*) before the study. Patients who get diagnosed, die of breast cancer or of other causes before the beginning of the study are not included in the simulated population. A time step of 1 day is chosen for numerical simulation. Due to the nature of discretization, possibilities of two events occurring during a period of a time step are encompassed in our numerical simulations. In particular, Algorithm I in [29] was used to speed up the simulation of Stage 1 and also eliminate the simultaneous occurrence of undetected cancer and non-breast cancer death events. Because implementation of Algorithm I requires the integration of the cancer incidence rate *c*_*a*_(*t*), we used a polynomial approximation of *c*_*a*_(*t*). Moreover, we considered all possible cases including the concurrence of detected cancer and non-breast cancer death events when simulating Stage 2. At any time during the follow-up, if the death event occurs, the simulation is terminated.

Let *X*_*i*_ be the simulated outcomes, the number of detected cancers and the number of cancer deaths, and *Y*_*i*_ be the corresponding recorded observations from the study. For the *j*th realization of our model, *S*_*j*_ is the sum of squared deviations, i.e. *S*_*j*_ =Σ_*i*_(*X*_*i*_ − *Y*_*i*_)^2^. The three unknown parameters (*b*_*b*2_, *γ* and *Z*) are chosen to minimize the expected value of *S*. Model calibration is carried out using the data only from the control arm. Then we simulate the model with the estimated parameters over both arms to reproduce the outcomes of the CNBSS.

### 2.4. Statistics and calculation of overdiagnosis

To quantify the survival and overdiagnosis, we simulated each patient in the BCHAM model 100 times, 50 times in the mammography and 50 in the control arm, with identical parameters and time of onset of cancer. We used Cox proportional hazards (the coxph function in R [30]) to evaluate the effect of treatment arm on survival from the time of acquiring cancer, thus avoiding the effects of lead time bias [18]. To illustrate the effects, we conduct these regressions on data broken up into sextiles of aggressiveness *k* and maximum tumor diameter *d*_*max*_, and by the time of cancer acquisition before, during or after the study. We term these 108 groups as study subcohorts. To estimate confidence limits, we bootstrap the simulated patients by sampling with replacement.

We quantify the number of diagnoses by computing the number of patients diagnosed in each arm and comparing in each subcohort with a *χ*^2^ test. To test for overdiagnosis itself, we compute the probability of death from other causes before death from cancer using the hazards *h* and *h*_*a*_ from Table 1a and integrating as in [31]. To minimize confounding the benefits of treatment that sufficiently delay cancer-induced mortality to allow death from other causes, we lower the treatment effectiveness parameter in the cancer mortality hazard *h* to its minimum value *α* = 2.5.

## 3. Results

### 3.1. Fit with CNBSS results

Simulations of BCHAM accurately capture the outcomes of the CNBSS including the number of detected cancers, deaths from cancer, deaths from other causes and the distributions of age at diagnosis in both arms (Figures 2a, 2b and Table 2).

**Table 2.** Comparison of simulated versus recorded ages at diagnosis (at cancer death in 25 years) for breast cancer detecting during screening phase (from the beginning of follow-up to the 5^th^ year) in mammography arm versus control arm.

|  | Mean | Range |
| --- | --- | --- |
|  | Simulated (recorded) | Simulated (recorded) |
| Age at diagnosis (years): |  |  |
| In MA | 53.08 (52.5) | 40.00 – 63.84 (40 – 64) |
| In CA | 53.44 (52.6) | 40.02 – 63.82 (40 – 64) |
| Age at cancer death (years): |  |  |
| In MA | 60.97 (59.9) | 40.56 – 80.76 (43 – 80) |
| In CA | 60.70 (60.6) | 41.43 – 80.30 (43 – 83) |

**Figure 2.**
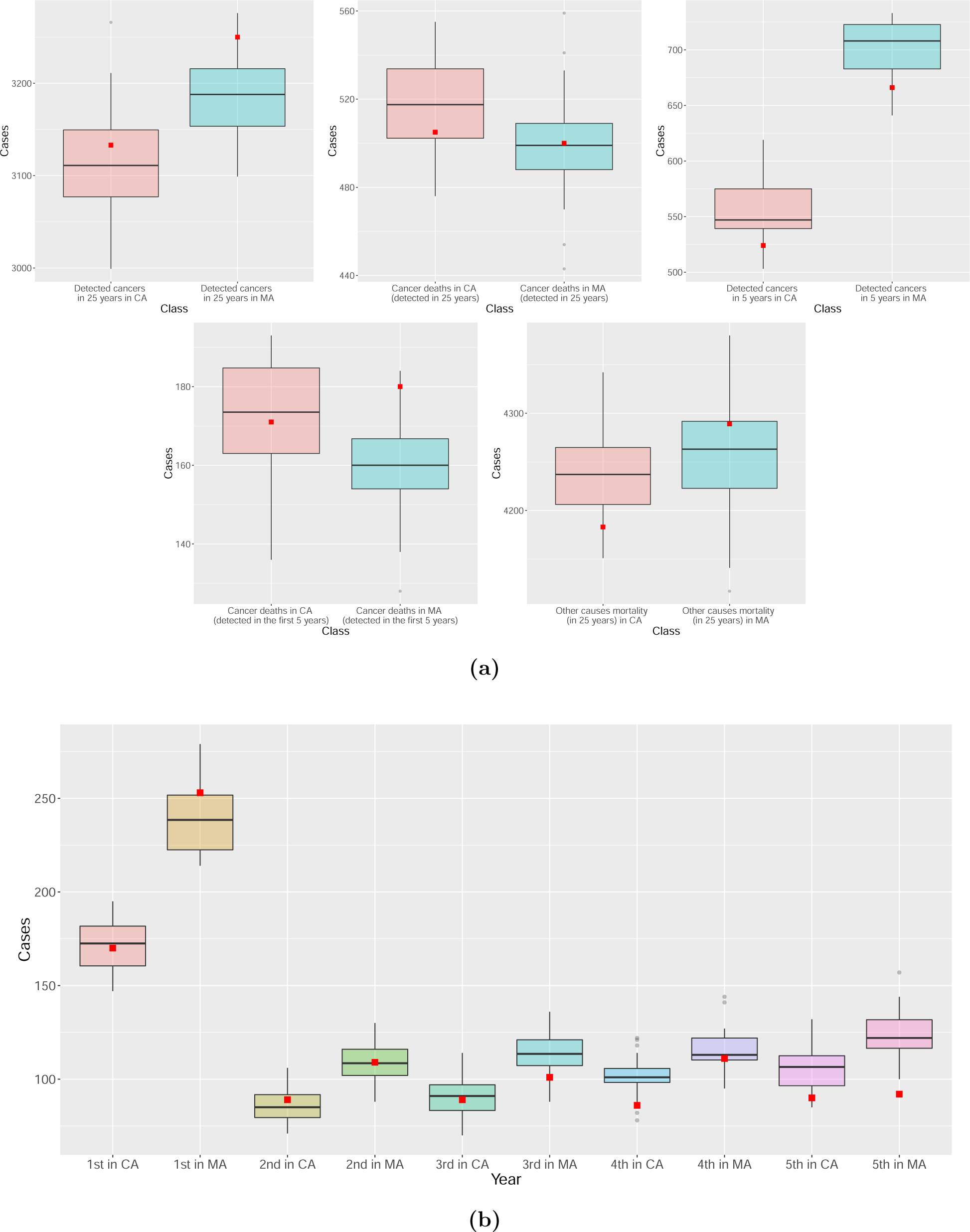
a) The number of simulated (box-plots) versus recorded (▪, CNBSS) breast cancers diagnosed and deaths from breast cancer in mammography arm (MA) and control arm (CA). b) Simulated (box-plots) versus recorded (▪, CNBSS) number of breast cancers diagnosed in MA and CA by study year.

### 3.2. Benefit and harm of mammography screening

The accurate fit of the model to the data under current conditions motivates testing how various model parameters affect the balance between benefit and harm. We first quantify the benefit of increasing the parameter *α* that describes the effectiveness of treatment (Figure 3a). Value of screening is estimated as the percent increase in survival after 25 years of follow-up. As can be seen, when cancer treatment is highly effective, mammography screening provides only a minimal benefit.

**Figure 3.**
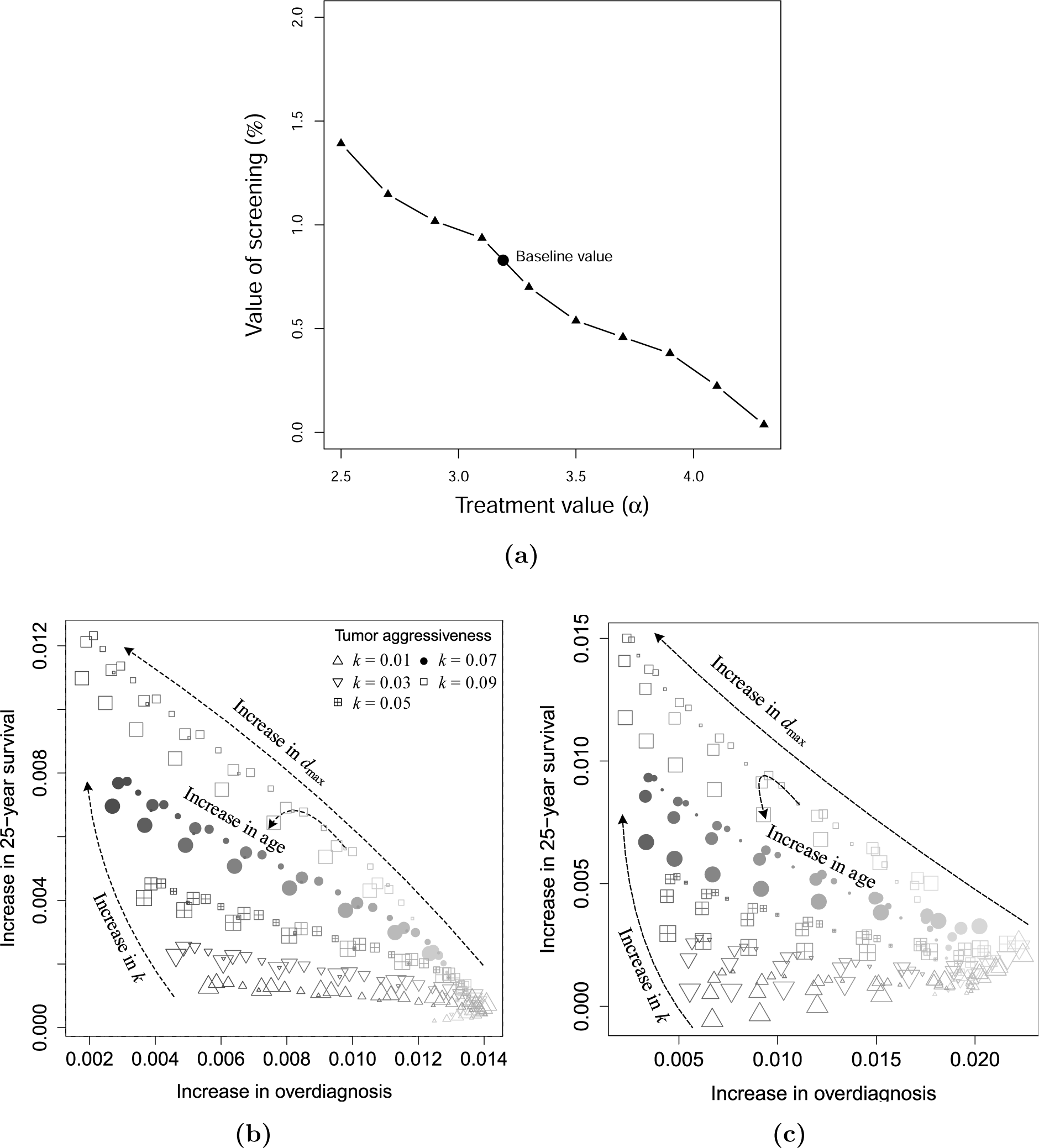
a) Survivorship as a function of the treatment effectiveness parameter *α*. b) Benefit (increase in probability of surviving patients after 25 years of follow-up) and harm (increase in probability of patients diagnosed with cancers that would not have been the cause of death) of mammography screening. c) Benefit and harm of mammography screening with an increase of breast cancer incidence by a factor of 5 in comparison with the baseline case presented on the left. Size of dots indicates age, the size increases with age. Color saturation increases with value of *d*_*max*_. Markers indicate value of *k*, △ (□) corresponds to the smallest (largest) value of *k*.

To compare the benefit and harm as a function of age, tumor aggressiveness *k*, and maximum tumor diameter *d*_*max*_, we simulate identical populations in both arms. In the simulation, participants receive an annual mammography and physical examinations in the mammography arm, or only an annual physical examination in the control for the first five years of follow-up. The simulation of each patient was repeated 50 times to reduce variance due to individual variation. Higher values of *k* and *d*_*max*_ strongly increase survival and decrease overdiagnosis (Figure 3b). The effects of age are much weaker, with slightly improved benefits in the middle age groups (women between the ages of 44 and 56). For patients with unaggressive tumors (small values of *k* and/or *d*_*max*_), mammography provides little benefit and the highest harm.

We also illustrate overdiagnosis and survivorship as functions of whether patients first acquired their cancer before, during, or after the study, and of the aggressiveness (*k*) and the maximum tumor diameter *d*_*max*_ of their tumor. For overdiagnosis (Figures 4a and b, we compare the difference in number of patients per thousand diagnosed in the mammography and control arms (top number) with the difference in the number of patients per thousand who would have died of other causes with less effective treatment (*α* = 2.5, bottom number). For survivorship (Figures 4c and d, we compare the difference in number of deaths per thousand in the mammography and control arms (top number) with the hazard ratio of death due to inclusion in the mammography arm (bottom number).

**Figure 4.**
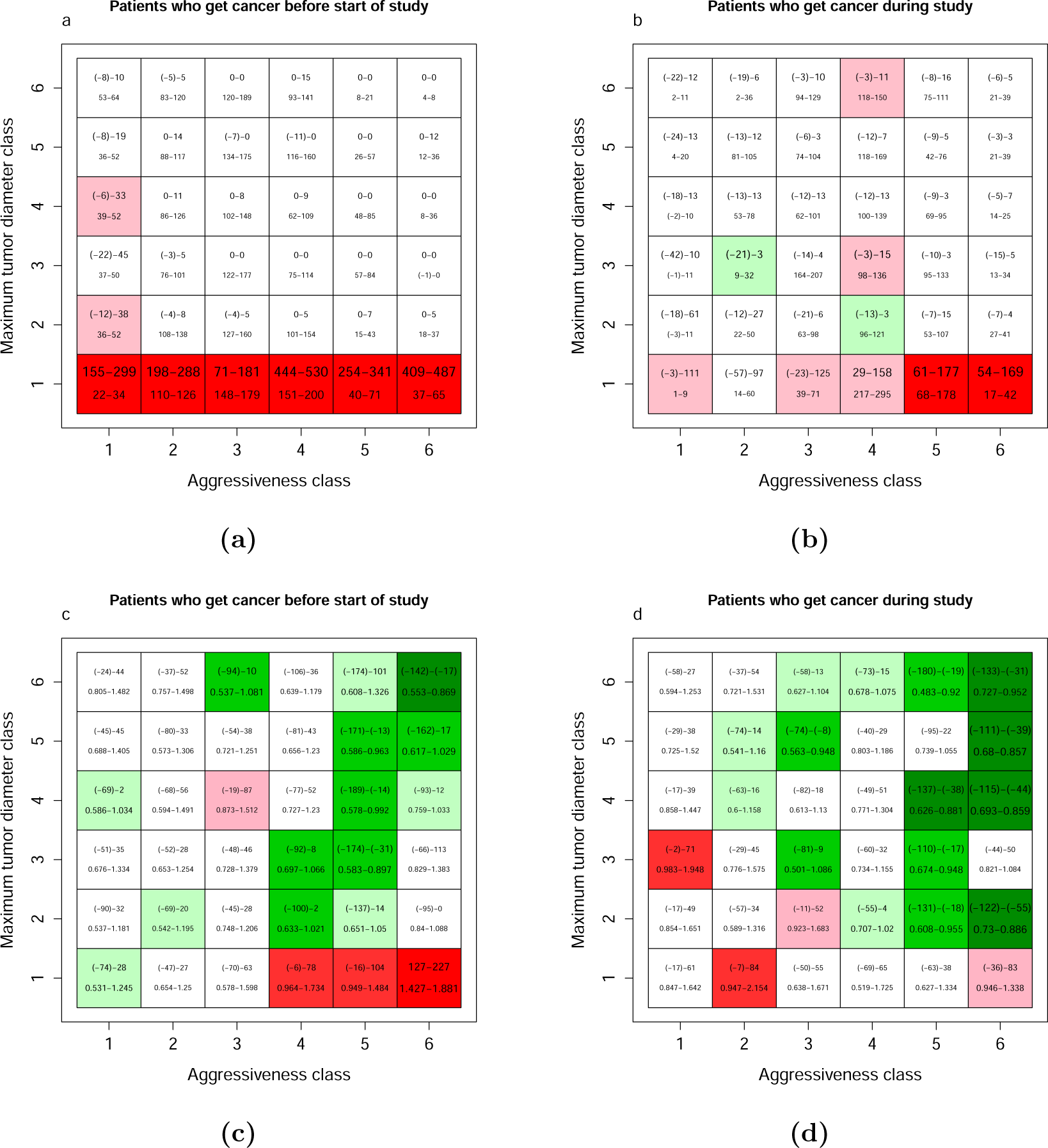
Comparison of simulated mammography and control arms after bootstrap analysis. The aggressiveness class indicates the value of the tumor aggressiveness parameter *k* (1 represents *k* < 0.0275, 2 from 0.0275 − 0.0400, 3 from 0.040 − 0.0543, 4 from 0.0543 − 0.0726, 5 from 0.0726 − 0.104 and 6 values greater than 0.104). The maximum tumor diameter class indicates the parameter *d*_*max*_, with all values in mm (1 represents *d*_*max*_ < 22.36, 2 from 22.36 − 43.36, 3 from 43.36 − 64.16, 4 from 64.16 − 85.05, 5 from 85.05 − 106.4 and 6 values greater than 106.4). Ranges are from 500 bootstrap replicates of the simulated data. Panels a) and b) show the difference in the number of patients per thousand diagnosed (top number) and the difference in the number per thousand who would have died first of other causes with ineffective treatment (bottom number, *α* = 2.5). Colors indicate the significance of the difference in probability of diagnosis in the two arms (red for higher in mammography arm, green for lower). Panels c) and d) show the difference in number of patients per thousand who died of any cause (top number) and the hazard ratio associated with mammography (bottom number) and colors indicate the significance of the effect of mammography on survival (red for higher hazard in the mammography arm, green for lower).

For patients who acquired an undetected tumor before the beginning of the study, those with ultimately small tumors are highly overdiagnosed and experience reduced survival due to the effects of the treatment. The greatest survival benefits accrue to patients with largest and most aggressive tumors. Patients who acquire a tumor during the five years of the study show a similar but weaker pattern of overdiagnosis, and no strong survival cost of overtreatment of rapidly-growing but ultimately small tumors. Patients who acquired tumors after the conclusion of the study show no effect of mammography as expected (results not shown). These observations suggest that overdiagnosis mainly occurs at the first screening [32].

To capture a high-risk population, such as women with germline BRCA1 and BRCA2 mutations, family breast cancer history, hormone therapy and smoking history, we assume an increase of breast cancer incidence by a factor of 5 [33] over the baseline case. The main observations remain largely unchanged. In a high risk population, mammography screening is slightly more beneficial for younger women, illustrated by a slight shift in the age effect in Figure 3c compared with Figure 3b.

## 4. Discussion

We have developed an individual-based mechanistic model of breast cancer incidence and mortality in a population based on the Canadian National Breast Screening Study (CNBSS). All but three of the parameters could be estimated independently from the literature or taken from the CNBSS report, with the remaining ones calibrated to the outcomes of the CNBSS. The model includes two forms of heterogeneity: tumor aggressiveness describing the growth rate and maximum tumor size.

The model accurately matches the cancer incidence and survival in the CNBSS (Section 3). We then use the model to quantify the benefit and harm of mammography screening, with benefit measured as the increase in 25-year survival, and harm as the increase in overdiagnosis. The benefit of screening decreases almost to zero with highly effective treatment. In general, patients with the most rapidly growing or potentially largest tumors have the highest benefit and least harm from mammography screening, with only a relatively small effect of age.

We measured overdiagnosis in two ways, through the difference in the number of patients diagnosed (excess incidence [18]), and through the number who would have died of other causes if treatment were relatively ineffective. The goal of treatment is, of course, to ensure that all patients have the chance to die of something else, and thus comparing the number of deaths with relatively effective treatment confounds true overdiagnosis with successful treatment. An alternative defines overdiagnosis as cancers that would not have presented clinically during the patient’s lifetime [15,31] which is most appropriate for modeling studies that optimize timing and type of testing.

Unlike age or other know risk factors, it is difficult in practice to predict specific tumor characteristics in an individual patient before recommending screening. In addition to improving mammography technology, increasing the net benefit of screening may require pretreatment tests that can identify women at the greatest risk of the highly aggressive cancers.

The CNBSS has been criticized because participants were volunteers [34] and thus possibly at higher risk than the general population. However, because participants were then randomized, this selection of volunteers should only increase the effect size of screening, but not create a change in direction.

Our model has several limitation. The parameters from the literature come from a variety of sources and studies that might not apply across all populations. The remaining three are based on a single study, and future work will test how effectively it can reproduce the outcome of other clinical trials, such as the Swedish trial [6,7], by modifying few or no parameter values. In addition, our modeling of treatment is quite simplified, without taking into account recent improvements or different treatments for different breast cancer types.

Our model brings a new quantitative tool to bear on the controversy over the use of mammography screening. We have found, in line with recent trials, that the benefits are sufficiently small and the harm sufficiently large to make screening of dubious value except in patients destined to have highly aggressive cancers, who of course are difficult if not impossible to identify in advance.

## Data Availability

NA

## Acknowledgment

We thank Dr. Adam Cohen for useful comments on an earlier draft, and the members of sLaM for discussion.

## Author contributions

All the authors contributed equally to the manuscript.

## Additional information

### Ethics approval and consent to participate

Ethical approval and consent were not required since this work was conducted based on published data in the literature.

## References

[1] R. L. Siegel, K. D. Miller, and A. Jemal, “Cancer statistics, 2018,” CA: A Cancer Journal for Clinicians, vol. 68, no. 1, pp. 7–30, 2018.

[2] S. Shapiro, P. Strax, and L. Venet, “Periodic breast cancer screening in reducing mortality from breast cancer,” Jama, vol. 215, no. 11, pp. 1777–1785, 1971.

[3] M. Roberts, F. Alexander, T. Anderson, A. Forrest, W. Hepburn, A. Huggins, A. Kirkpatrick, J. Lamb, W. Lutz, and B. Muir, “The Edinburgh randomised trial of screening for breast cancer: description of method,” British journal of cancer, vol. 50, no. 1, p. 1, 1984.

[4] F. Alexander, T. Anderson, H. Brown, A. Forrest, W. Hepburn, A. Kirkpatrick, C. McDonald, B. Muir, R. Prescott, S. Shepherd, et al., “The Edinburgh randomised trial of breast cancer screening: results after 10 years of follow-up,” British journal of cancer, vol. 70, no. 3, p. 542 1994.

[5] A. B. Miller, C. Wall, C. J. Baines, P. Sun, T. To, and S. A. Narod, “Twenty five year follow-up for breast cancer incidence and mortality of the Canadian National Breast Screening study: randomised screening trial,” Bmj, vol. 348, p. g366, 2014.

[6] S. W. Duffy, L. Tabar, B. Vitak, M. Yen, J. Warwick, R. Smith, and H. Chen, “The Swedish Two-County Trial of mammographic screening: cluster randomisation and end point evaluation,” Annals of Oncology, vol. 14, no. 8, pp. 1196–1198, 2003.

[7] L. Tabár, B. Vitak, T. H.-H. Chen, A. M.-F. Yen, A. Cohen, T. Tot, S. Y.-H. Chiu, S. L.-S. Chen, J. C.-Y. Fann, J. Rosell, et al., “Swedish two-county trial: impact of mammographic screening on breast cancer mortality during 3 decades,” Radiology, vol. 260, no. 3, pp. 658–663, 2011.

[8] R. A. Smith, S. W. Duffy, R. Gabe, L. Tabar, A. M. Yen, and T. H. Chen, “The randomized trials of breast cancer screening: what have we learned?,” Radiologic Clinics, vol. 42, no. 5, pp. 793–806, 2004.

[9] A. Bleyer and H. G. Welch, “Effect of three decades of screening mammography on breast-cancer incidence,” New England Journal of Medicine, vol. 367, no. 21, pp. 1998–2005, 2012.

[10] H. G. Welch, P. C. Prorok, A. J. O’Malley, and B. S. Kramer, “Breast-cancer tumor size, overdiagnosis, and mammography screening effectiveness,” New England Journal of Medicine, vol. 375, no. 15, pp. 1438–1447, 2016.

[11] H. G. Welch and E. S. Fisher, “Income and cancer overdiagnosis–when too much care is harmful,” New England Journal of Medicine, vol. 376, no. 23, pp. 2208–2209, 2017. PMID: 28591536.

[12] L. J. Esserman, “The WISDOM Study: breaking the deadlock in the breast cancer screening debate,” NPJ breast cancer, vol. 3, no. 1, p. 34 2017.

[13] M. Tsunematsu and M. Kakehashi, “An analysis of mass screening strategies using a mathematical model: comparison of breast cancer screening in Japan and the United States,” Journal of epidemiology, vol. 25, no. 2, pp. 162–171, 2015.

[14] S. Molani, M. Madadi, and W. Wilkes, “A partially observable Markov chain framework to estimate overdiagnosis risk in breast cancer screening: Incorporating uncertainty in patients adherence behaviors,” Omega, vol. 89, pp. 40–53, 2019.

[15] N. Gunsoy, M. Garcia-Closas, and S. Moss, “Estimating breast cancer mortality reduction and overdiagnosis due to screening for different strategies in the United Kingdom,” British journal of cancer, vol. 110, no. 10, p. 2412 2014.

[16] Y. Chen, G. Brock, and D. Wu, “Estimating key parameters in periodic breast cancer screening-application to the Canadian National Breast Screening Study data,” Cancer epidemiology, vol. 34, no. 4, pp. 429–433, 2010.

[17] S. Taghipour, D. Banjevic, A. Miller, N. Montgomery, A. Jardine, and B. Harvey, “Parameter estimates for invasive breast cancer progression in the Canadian National Breast Screening Study,” British journal of cancer, vol. 108, no. 3, p. 542 2013.

[18] R. De Gelder, E. A. Heijnsdijk, N. T. Van Ravesteyn, J. Fracheboud, G. Draisma, and H. J. De Koning, “Interpreting overdiagnosis estimates in population-based mammography screening,” Epidemiologic reviews, vol. 33, no. 1, pp. 111–121, 2011.

[19] D. Shaevitch, S. Taghipour, A. B. Miller, N. Montgomery, B. Harvey, et al., “Tumor size distribution of invasive breast cancers and the sensitivity of screening methods in the Canadian National Breast Screening Study,” Journal of cancer research and therapeutics, vol. 13, no. 3, p. 562 2017.

[20] L. A. Gaudette, C. Silberberger, C. A. Altmayer, and R.-N. Gao, “Trends in breast cancer incidence and mortality,” 1996.

[21] “Statistics Canada. Table 13-10-0710-01 Deaths and mortality rates, by age group,” 1991.

[22] L. Norton, “A Gompertzian model of human breast cancer growth,” Cancer research, vol. 48, no. 24 Part 1, pp. 7067–7071, 1988.

[23] J. A. Spratt, D. Von Fournier, J. S. Spratt, and E. E. Weber, “Decelerating growth and human breast cancer,” Cancer, vol. 71, no. 6, pp. 2013–2019, 1993.

[24] H. Weedon-Fekjær, S. Tretli, and O. O. Aalen, “Estimating screening test sensitivity and tumour progression using tumour size and time since previous screening,” Statistical methods in medical research, vol. 19, no. 5, pp. 507–527, 2010.

[25] J. S. Michaelson, L. L. Chen, M. J. Silverstein, M. C. Mihm Jr, A. J. Sober, K. K. Tanabe, B. L. Smith, and J. Younger, “How cancer at the primary site and in the lymph nodes contributes to the risk of cancer death,” Cancer: Interdisciplinary International Journal of the American Cancer Society, vol. 115, no. 21, pp. 5095–5107, 2009.

[26] J. S. Michaelson, M. Silverstein, J. Wyatt, G. Weber, R. Moore, E. Halpern, D. B. Kopans, and K. Hughes, “Predicting the survival of patients with breast carcinoma using tumor size,” Cancer, vol. 95, no. 4, pp. 713–723, 2002.

[27] A. S. Wahed, T. M. Luong, and J.-H. Jeong, “A new generalization of Weibull distribution with application to a breast cancer data set,” Statistics in medicine, vol. 28, no. 16, pp. 2077–2094, 2009.

[28] H. Weedon-Fekjær, B. H. Lindqvist, L. J. Vatten, O. O. Aalen, and S. Tretli, “Breast cancer tumor growth estimated through mammography screening data,” Breast Cancer Research, vol. 10, no. 3, p. R41, 2008.

[29] V. H. Thanh and C. Priami, “Simulation of biochemical reactions with time-dependent rates by the rejection-based algorithm,” The Journal of chemical physics, vol. 143, no. 5, p. 08B601_1, 2015.

[30] T. M. Therneau, A Package for Survival Analysis in S, 2015. version 2.38.

[31] M. Madadi, M. Heydari, S. Zhang, E. Pohl, C. Rainwater, and D. L. Williams, “Analyzing overdiagnosis risk in cancer screening: A case of screening mammography for breast cancer,” IISE Transactions on Healthcare Systems Engineering, vol. 8, no. 1, pp. 2–20, 2018.

[32] S. W. Duffy, O. Agbaje, L. Tabar, B. Vitak, N. Bjurstam, L. Björneld, J. P. Myles, and J. Warwick, “Overdiagnosis and overtreatment of breast cancer: estimates of overdiagnosis from two trials of mammographic screening for breast cancer,” Breast Cancer Research, vol. 7, no. 6, p. 258 2005.

[33] A. W. Kurian, G. D. Gong, E. M. John, D. A. Johnston, A. Felberg, D. W. West, A. Miron, I. L. Andrulis, J. L. Hopper, J. A. Knight, et al., “Breast cancer risk for noncarriers of family-specific BRCA1 and BRCA2 mutations: findings from the Breast Cancer Family Registry,” Journal of Clinical Oncology, vol. 29, no. 34, p. 4505 2011.

[34] A. Miller, G. Howe, and C. Wall, “The national study of breast cancer screening,” Clin Invest Med, vol. 4, pp. 227–258, 1981.

